# Prediction of the time evolution of the COVID-19 disease in Guadeloupe with a stochastic evolutionary model

**DOI:** 10.1101/2020.04.12.20063008

**Authors:** Meriem Allali, Patrick Portecop, Michel Carlès, Dominique Gibert

**Affiliations:** Emergency Medical Service (SAMU-SAU), University Hospital, Pointe à Pitre, Guadeloupe; Critical Care Department, University Hospital, Pointe à Pitre, Guadeloupe; Univ Lyon, Univ Lyon 1, ENSL, CNRS, UMS 3721, LGL-TPE, F-69622 Villeurbanne, France

**Keywords:** COVID-19, Time-evolution, Guadeloupe, SAMU, Critical care, Monte Carlo Model

## Abstract

Predictions on the time-evolution of the number of severe and critical cases of COVID-19 patients in Guadeloupe are presented. A stochastic model is purposely developed to explicitly account for the entire population (≃400000 inhabitants) of Guadeloupe. The available data for Guadeloupe are analysed and combined with general characteristics of the COVID-19 to constrain the parameters of the model. The time-evolution of the number of cases follows the well-known exponential-like model observed at the very beginning of a pandemic outbreak. The exponential growth of the number of infected individuals is controlled by the so-called basic reproductive number, *R*_0_, defined as the likely number of additional cases generated by a single infectious case during its infectious period *T*_*I*_. Because of the rather long duration of infectious period (≃14 days) a high rate of contamination is sustained during several weeks after the beginning of the containment period. This may constitute a source of discouragement for people restrained to respect strict containment rules. It is then unlikely that, during the containment period, *R*_0_ falls to zero. Fortunately, our models shows that the containment effects are not much sensitive to the exact value of *R*_0_ provided we have *R*_0_ *<* 0.6. For such conditions, we show that the number of severe and critical cases is highly tempered about 4 to 6 weeks after the beginning of the containment. Also, the maximum number of critical cases (i.e. the cases that may exceed the hospital’s intensive care capacity) remains near 30 when *R*_0_ *<* 0.6. For a larger *R*_0_ = 0.8 a slower decrease of the number of critical cases occurs, leading to a larger number of deceased patients. This last example illustrates the great importance to maintain an as low as possible *R*_0_ during and after the containment period. The rather long delay between the beginning of the containment and the appearance of the slowing-down of the rate of contamination puts a particular strength on the communication and sanitary education of people. To be mostly efficient, this communication must be done by a locally recognised medical staff. We believe that this point is a crucial matter of success. Appendix Posterior model assessment with data acquired after April 11, 2020 added in a second version of the paper compares the model predictions with the data acquired from April 12 to May 25 2020, after the construction of the model discussed in the present study. The remarkable agreement between the model predictions and the data may be explained by the good quality of first-hand data used to constrain the model, the ability of the stochastic approach to integrate new information and stability of the sanitary situation due to the respect of the recommendations emitted by medical and administrative authorities by the guadeloupean population.

## Introduction

The most recent evolution of the pandemic COVID-19 disease in Western Europe indicates that this region is, together with the United States, the new centre of the pandemic spread (e.g. (1) and other reports issued by the World Health Organization). Italy and Spain are confronted with large outbreaks of SARS-CoV-2 infection. In France, the rate of new infections daily increases and measures have already been taken to increase the intensive care capacity of the main hospitals of the country. Also, in order to face with the strong heterogeneity of case number among the different regions in France, medevacs (either by air or railway) have been undertaken to optimally redistribute the most critical patients in the country’s intensive care facilities. In this context, the situation of remote French territories like Guadeloupe is particularly critical since, although possible, medevacs should be anticipated with a longer delay because of the distance and the duration of the travels.

Numerical models of the spread of epidemic diseases may be of some help to anticipate the evolution of the situation in a near-future of several weeks and, eventually, may reveal a likely disruption of the local intensive care capacity. In short, mathematical models may be ranked in two main categories, namely semi-analytical models and numerical stochastic and Monte Carlo models (see (2) for a review). In the former category, the spread of the disease is modelled by a set of coupled differential equations that account for the most important characteristics of the disease. This approach is largely followed (3–5). The second category of models is, in some sense, more straightforward and relies on network models to explicitly considers the individuals constituting the population. Such an approach offers a great versatility to tackle with complicated features, like social interaction matrices, that are difficult to introduce in semi-analytical models. The main drawback of numerical stochastic models is their computer-intensive demand that, for large populations, necessitates the use of multi-scale or coarse-grained algorithms. Thanks to the moderate size of the population of Guadeloupe, no such difficulties are encountered and a straightforward approach is possible.

## Method

The technical details of the model are explained in the appendix Stochastic Monte Carlo model, and we here recall its main characteristics. A flowchart of the model is shown in Figure 1. As stated above, all individuals forming the population are considered as nodes in a fully connected network where everyone is able to meat anyone. By using social contact matrices, this full connection could be modified to account for demographic and social heterogeneity. Also, we have not considered the age-dependence of the COVID-19 effects.

**Fig. 1.**
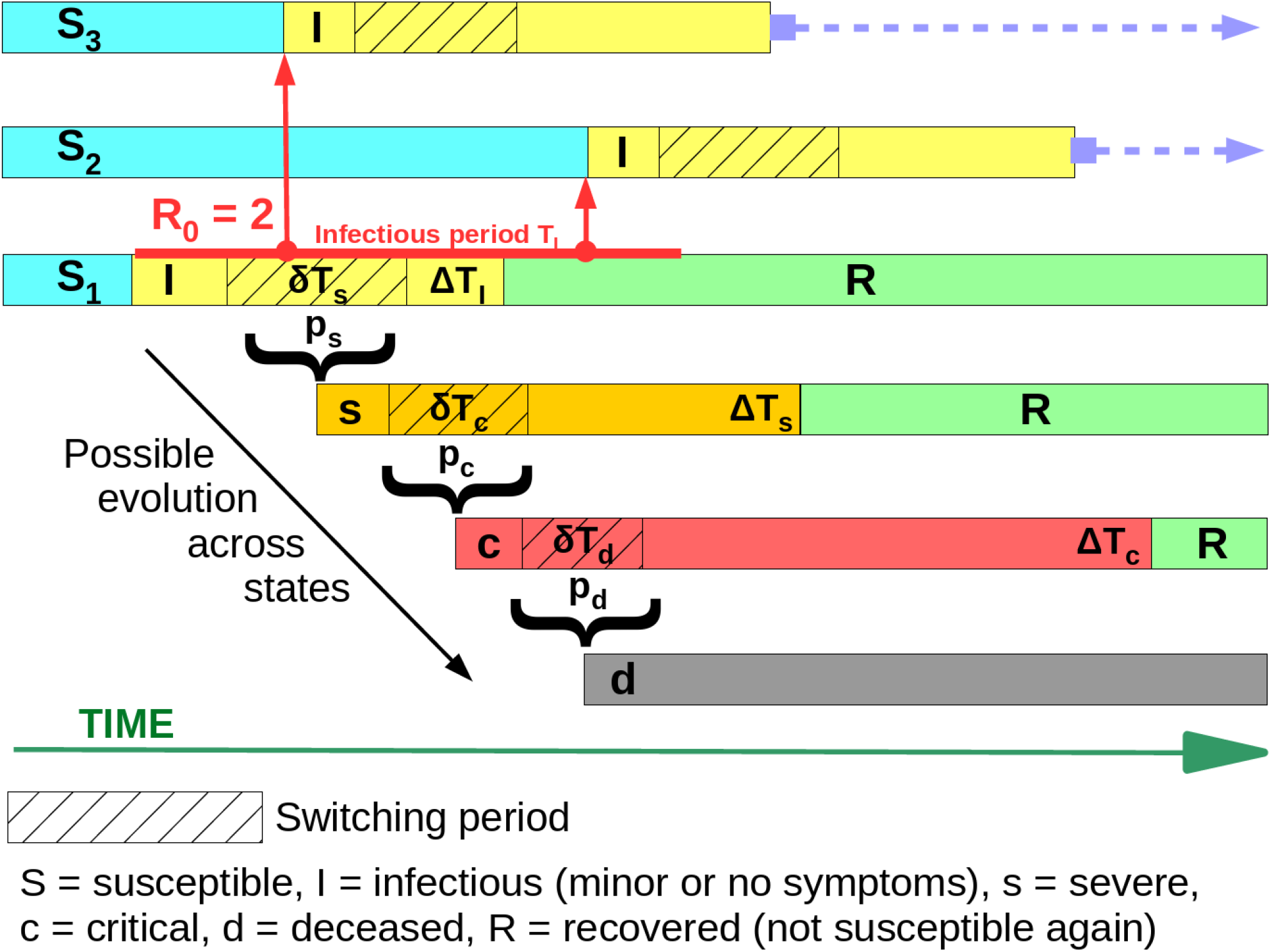
Flowchart of the stochastic modelling procedure. In the general case, a susceptible non-infected person *S*_1_ becomes infected. This new infected *I* may contaminate a number *R*_0_ of other susceptibles (here *S*_2_ and *S*_3_) during his infected period *T*_*I*_ (red line) which may run beyond the recovery period Δ*T*_*I*_ (in yellow). During the sub-period *δT*_*s*_ (shaded rectangle), the infectious “I” may switch to state “severe” with a probability *p*_*s*_. If the patient remains in state “I” until the end of the recovery period Δ*T*_*I*_, he becomes definitively recovered “R”. Instead, if the patient switches to state “s”, he may either recover at the end of the recovery period Δ*T*_*s*_ or switch with a probability *p*_*c*_ to state critical “c” during the switching period *δT*_*c*_. The same procedure applies to state critical “c”.

Each individual of the network may, temporarily or definitely, be in the following state (Fig. 1): non-infected, infected with minor symptoms (“infectious”), infected with severe symptoms (“severe”), infected critical (“critical”), dead or recovered. In the vocabulary of epidemic modelling, non-infected correspond to the so-called “Susceptibles” and minor infected are “Infectious”. In our model, both the severe and critical infected are not considered as infectious because they are isolated in hospital facilities and unable to significantly contaminate others. Although this is statistically justified in our model, actually this assumption is contradicted by the sad death of several French medics.

According to the classical nomenclature, our model is a SIscRd model where the lowercase “sc” indicate the transient and non-contaminating nature of these states. To the best of our knowledge at the time of writing this paper, it does not seem that recovered “R” patients are able to again become infectious “I” (6). The deceased “d” patients may remain infectious several days (7) and we assume that they are safely isolated to prevent any contamination.

Each individual may switch from one state to another with given probabilities. For instance, a “susceptible” may become “infectious”, then “severe” and finally “recovered”. This example corresponds to the sequence:

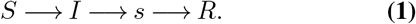

The duration of stay in a given class is variable, depending on the initial health status of each patient. Clinical data collected world-wide put constrains on the possible range of each parameter.

The model is based on an evolutionary scheme where the initial conditions correspond to a non-infected population excepted a small (typically several tens) number of “infectious”. Once initialised, the algorithm proceeds by time-steps and, for each time-step, the sequence of evolutionary operations is applied. For instance, for the time-step corresponding to day *k* of the simulation process:

1. All infectious, severe and critical patients that reached their respective recovery duration (i.e. Δ*T*_*I*_, Δ*T*_*s*_, Δ*T*_*c*_) are definitely switched to the state recovered “R” (Fig. 1).
2. The ensemble of infectious at day *k* may contaminate susceptibles “S” with a probability given by the *R*_0_ value at day *k*. By this way, the model is able to account for rapid time-changes of *R*_0_.
3. All infectious, severe and critical patients that are in their switching period (i.e. *δt*_*s*_, *δt*_*c*_ and *δt*_*d*_ in Fig. 1) may switch to the next stage with a given probability. This corresponds to the following possible transitions:

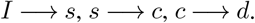

## Data

The data used in the present study, are daily communicated by the University Hospital to the local authorities, i.e. the Regional Health Agency (Agence Régionale de Santé in French). They correspond to the cumulative number of persons with COVID-19, the cumulative number of deceased patients and the number of patients presently in intensive care units. Detailed data for France are made available by Santé Publique France (8).

Both the cumulative number of deceased patients and the number of patients presently in intensive care units respectively correspond to Σ*N*_*d*_ and *N*_*c*_ in the model. The cumulative number of persons with COVID-19 could be something between Σ*N*_*I*_ and Σ*N*_*s*_, depending on the screening procedure. In France, a majority of the persons tested for COVID-19 are patients with severe symptoms and admitted in specialised COVID-19 units. Such is the case in Guadeloupe and, consequently, the cumulative number of persons with COVID-19 as announced by ARS correspond to the Σ*N*_*s*_ of the model.

In the present study, we use the data going from March 13 2020 to April 11 2020 shown in Figure 8 of appendix Boot-strapping method of data analysis to construct the predictive models discussed in the present study. In a second version of the present paper, the data set has been extended up to May 25 2020 to perform the posterior evaluation of the model performance presented in appendix Posterior model assessment with data acquired after April 11, 2020.

## Results

The model derived in the present study is highly non-linear with respect to most parameters, and it is expected that nonunique and significantly different solutions fitting the data might be obtained. This could be performed by the means of non-linear inverse methods like simulated annealing (9, 10) and will be presented in a forthcoming study. In the present study, the *Z*_*I*_ and time-varying *R*_0_ parameters are adjusted with the Nelder-Mead downhill simplex (11, 12). The other parameters are determined with clinical observations in the Guadeloupe hospital and data published in the abundant literature concerning COVID-19.

Figure 2 shows the results for model 1. The time variations of *R*_0_ have been adjusted to reproduce the flattening visible in the Σ*N*_*s*_ data from day 21. The corresponding values of the parameters are recalled in the upper-right panel of the Figure. This model provides a good fit to all data Σ*N*_*s*_, *N*_*c*_ and Σ*N*_*d*_. In order to reproduce the initial rapid exponential increase observed before day 10, quite large values of *R*_0_ = [4.2 4.2 4.2 4.5 4.5 4.5] are found for the first 6 days of the simulation (day 1 corresponds to March 11, 2020). These high *R*_0_ are obtained during the week before municipal elections when meetings occurred and were probably places of high contamination rates (13, 14). This could explain the high *R*_0_ values found with the model.

**Fig. 2.**
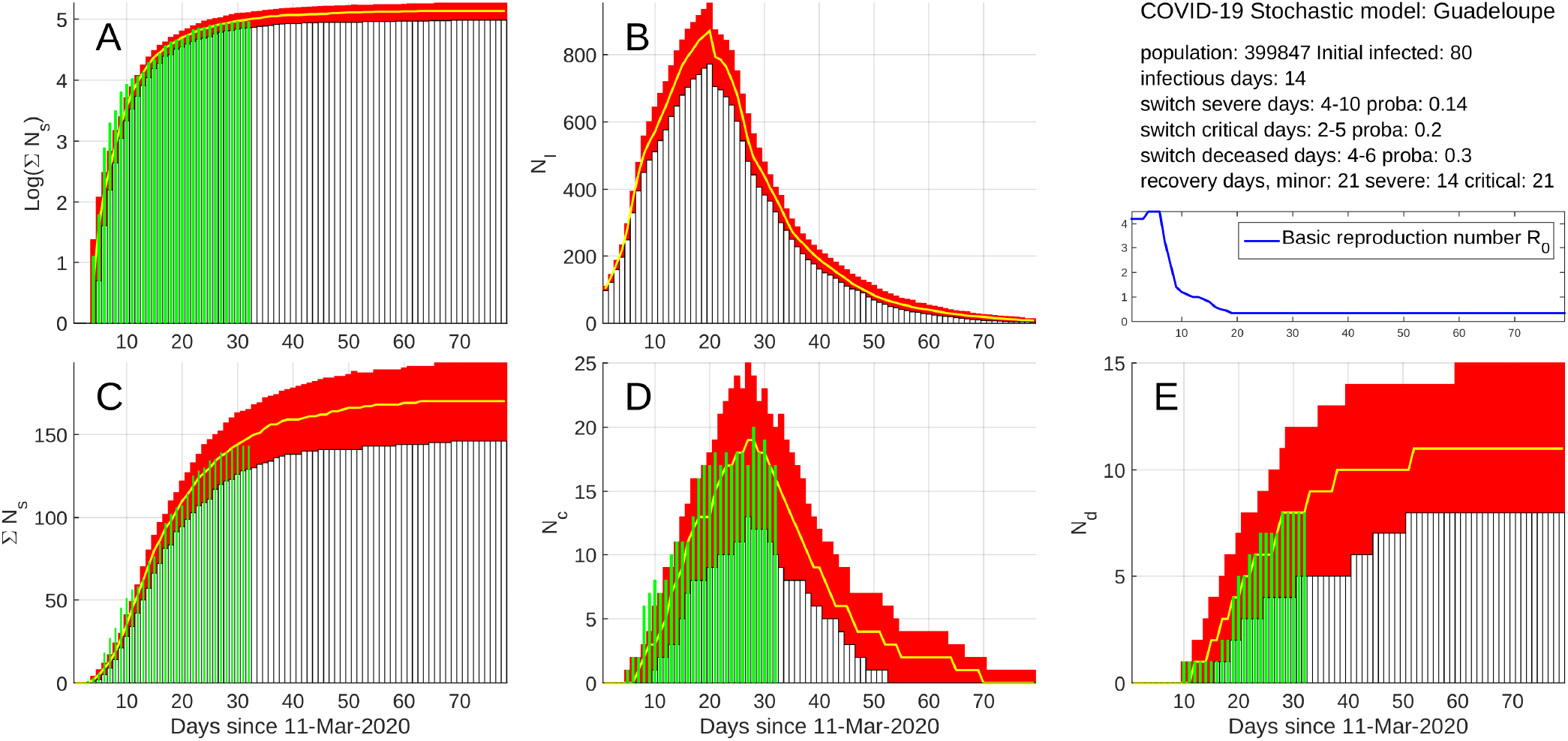
Model 1 results. A) semi-logarithmic (Natural logarithm) plot of the cumulative number Σ*N*_*s*_ of severe cases. Green bars = data and red bars = model. B) Instantaneous number *N*_*I*_ of infectious. C) same as (A) in linear axis. D) Instantaneous number *N*_*c*_ of critical cases. Green bars = data and red bars = model. E) Cumulative number Σ*N*_*d*_ of deceased patients. Green bars = data and red bars = model. The parameter values used in the model are shown in the upper-right part of the figure together with the time-variation of the basic reproductive number *R*_0_. The red rectangles represent the 80% confidence interval centred on the median.

Interestingly, the large *R*_0_ from days 1 to 6 must be combined with a large *Z*_*I*_ = 80 to fit the sharp a onset of the Σ*N*_*s*_ curve (Fig. 2A). The reasons for such a large number of initial infectious remain unknown, but we may suspect either a massive arrival of infected aircraft or ship passengers or the existence of several infectious spots like funeral wakes or election meetings as mentioned above.

In order to fit the strong inverse curvature of the *log*(Σ*N*_*s*_) curve between days 10 to 20, *R*_0_ must be gradually decreased from day 7 (March 17) to day 18 (March 28): *R*_0_ = [3.2 2.3 1.4 1.2 1.1 1.0 1.0 0.9 0.8 0.6 0.5 0.45]. The starting date of the decrease corresponds to the beginning of the containment following the second speech of the President of the French Republic, Mr Macron, on March 16. To reproduce the flat almost horizontal end of the data from day 21 (March 31), it is necessary to reduce *R*_0_ = 0.35 from day 19 (March 29) until the end of the process (i.e. day 80).

With this model, the maximum *N*_*c*_ = 25 *±* 3 is reached near day 28 (April 7) about 3 weeks after the beginning of the containment. After this date, the number of critical cases sharply decreases to reach a low base level about 4 weeks later, i.e. during the first week of May.

To illustrate the role played by *R*_0_ during the containment period, we present the results for 2 models with the same parameter values as for model 1 excepted during the containment. We set *R*_0_ = 0.6 in model 2 (Fig. 3) and *R*_0_ = 0.8 for model 3 (Fig. 4). Both models 2 and 3 fit the data as well as model 1 excepted for the flattening part of the Σ*N*_*s*_ data after day 21 (March 31). This indicates that the determination of *R*_0_ during the containment is constrained only by the most recent data values. The quality and reliability of these data is then of a great importance to derive models able to predict an eventual decrease of critical cases.

**Fig. 3.**
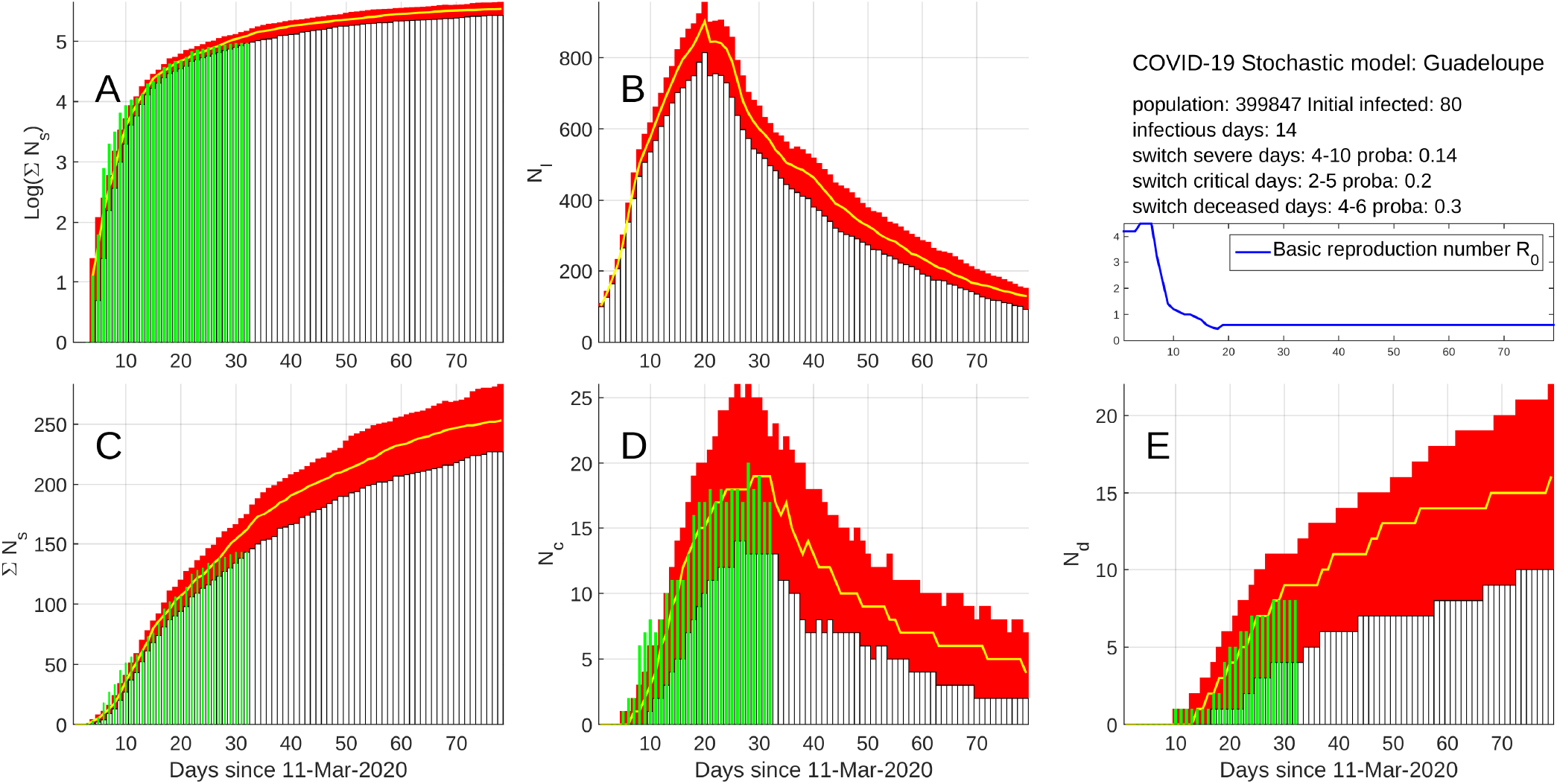
Results for model 2 with the same parameters as model 1 (Fig. 2) excepted for the containment *R*_0_ = 0.6 from day 19 (March 29).

**Fig. 4.**
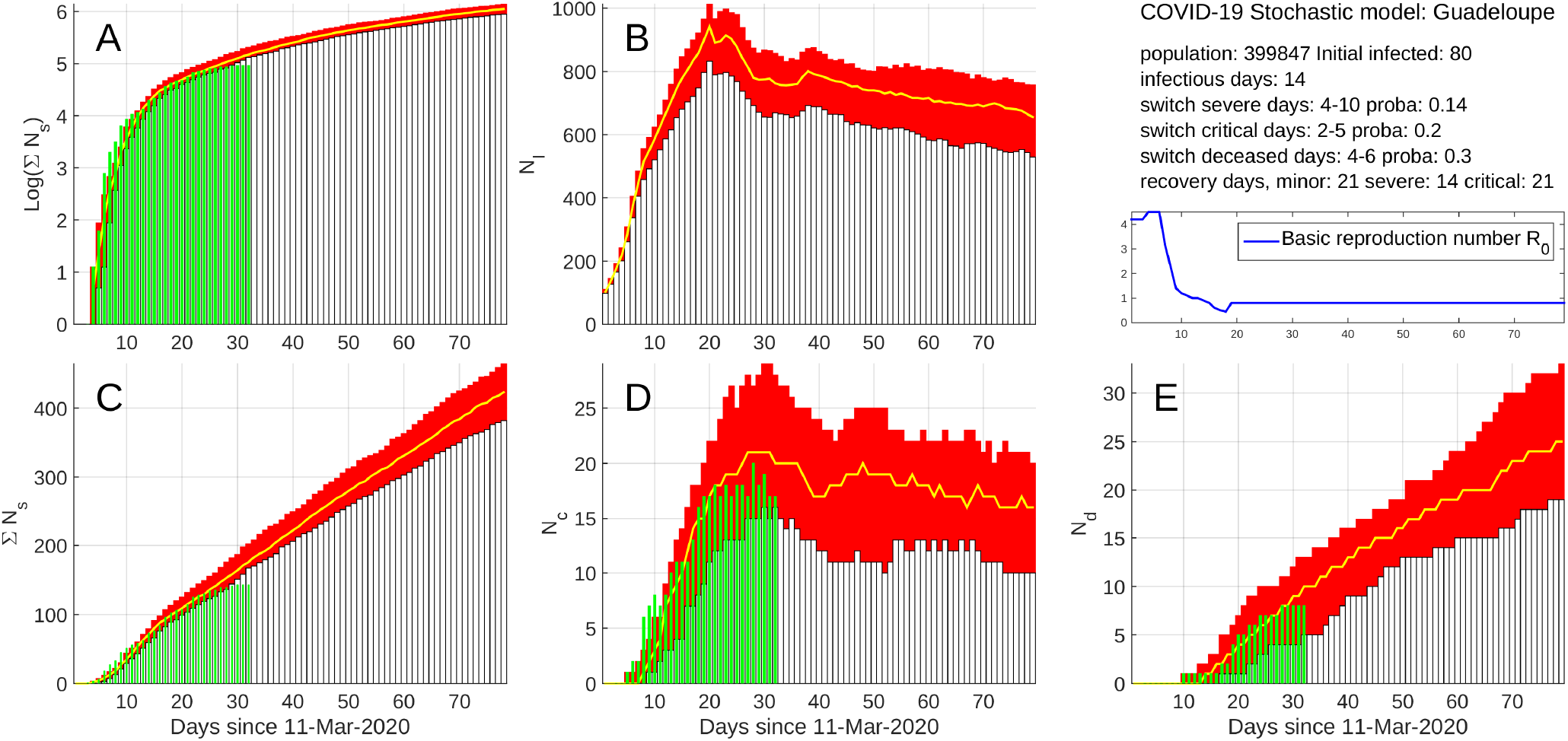
Results for model 3 with the same parameters as model 1 (Fig. 2) excepted for the containment *R*_0_ = 0.8 from day 19 (March 29).

Model 2 (Fig. 3) corresponds to the containment *R*_0_ = 0.6 and gives a maximum number of critical cases of the same order and at the same date as the one obtained with model 1 (Fig. 2). However, the decrease following the maximum is less steep and low values are reached about 3 weeks later with respect to what is observed with model 1. This translates into a larger cumulative number of critical case and, consequently, in a larger number of deceased patients (compare Fig. 2E and Fig. 3E).

Model 3 (Fig. 4) corresponds to *R*_0_ = 0.8. As can be verified in Figure 4A,C, the flattening of the data after day 21 (March 31) is poorly reproduced making this model less likely than models 1 and 2. However, owing that the flattening of the Σ*N*_*s*_ curve relies on a small part of the most recent data, this pessimistic model cannot be totally excluded at the time of writing this article. The maximum values of critical cases may reach a maximum of 30 critical patient followed by a plateau with a small slope during which the instantaneous number *N*_*c*_ remains around 15-20 one month after the date of the maximum. This correspond to a situation where the treatment of numerous critical patients must be sustained during a long period, implying the disposal of a sufficiently large medical staff and amount of equipment. As can be observed in Figure 4E the number of deceased patients increases steadily.

## Concluding remarks

A common characteristic to all 3 models presented above, is the need of a quite large number *Z*_*I*_ = 80 of initial infectious persons coupled with a large *R*_0_ ≃4 at the beginning of the epidemic spread. This large *Z*_*I*_ could be explained either by the importation of a large number of infected persons or the presence of several super contaminators able to contaminate tens of persons during meetings in a short period of time (see (13, 14) for the effects of mass gathering). Let us remark that large *R*_0_ are reported by others; for instance, Tang et al. report values as high as 6.47 for data from China. These authors mention that this high *R*_0_ corresponds to data collected during a period of intensive social contacts (i.e. before the Chinese New Year). Mizumoto and Chowell report *R*_0_ values as high as 10 for the case of Diamond Princess, and for the same data Rocklöv et al. find a maximum *R*_0_ = 14.8 and a 8-fold reduction to *R*_0_ = 1.78 during isolation and quarantine.

Another characteristic of the model is the need to significantly reduce *R*_0_ to fit the decelerating curvature of the Σ*N*_*I*_ data curve (e.g. Fig. 2A). This reduction is delayed by about 3 days with respect to the beginning of the containment and confirms an overall good respect of the social distancing rules by the population of Guadeloupe. Several French national media published articles stating that Guadeloupe was relativity spared from the disease (18). Such a claim could have triggered a common sense reflex of protection applied through social distancing and usage of rules of hygiene. To fit the most recent Σ*N*_*I*_ data, a low *R*_0_ = 0.35 must be applied. If true, this would indicate that people of Guadeloupe continued to improve their social behaviour during the 3 weeks after the beginning of the containment.

The models allows to get an estimate of the number of instantaneous infectious *N*_*I*_ and cumulative recovered patients Σ*N*_*R*_. In absence of systematic detection of COVID-19 among the population, no *N*_*I*_ data are available and the *N*_*I*_ curve is actually indirectly constrained by the fit to the Σ*N*_*s*_ and *N*_*c*_ data and by the switching probabilities *p*_*s*_ and *p*_*c*_. However, the values given to these probabilities fall in ranges widely recognised by the medical community and we may safely consider them sufficiently reliable to give credit to the modelled *N*_*I*_ and Σ*N*_*R*_ curves. A simple assessment may be done by dividing Σ*N*_*d*_ by the observed number of deceased patients. On day 28 (April 7), this ratio equals 0.7%, a value slightly lower than the generally recognised ratio of 1 − 2% (19).

For the models discussed above, several thousands of persons have been infected and a large fraction of them have recovered and are supposed protected against another infection. However, these supposedly protected persons represent a relatively small part of the total population and the number of susceptible persons remains sufficiently large to ensure a restart of a second epidemic spread of the disease. This is shown in Figure 5 which represents a long-term simulation with model 1 as in Figure 2 but with an abrupt resetting of *R*_0_ = 4.0 at day 70 (mid-May), about 2 months after the beginning of the containment. This corresponds to a situation of uncontrolled end of containment. Because of the existence of only several infectious cases, the spread of the virus proceed at a low-level during approximately 3 weeks (i.e. until day 90) before exponentially exploding again into a second epidemic crisis. These results illustrate the future difficulty to control such a restart of the virus propagation and the necessity to maintain a low *R*_0_ for a long period of time. The simulation shown in Figure 5 assumes that the patients who recovered during the first epidemic crisis cannot be infected during the second crisis, a medical assumption that remains to be confirmed.

**Fig. 5.**
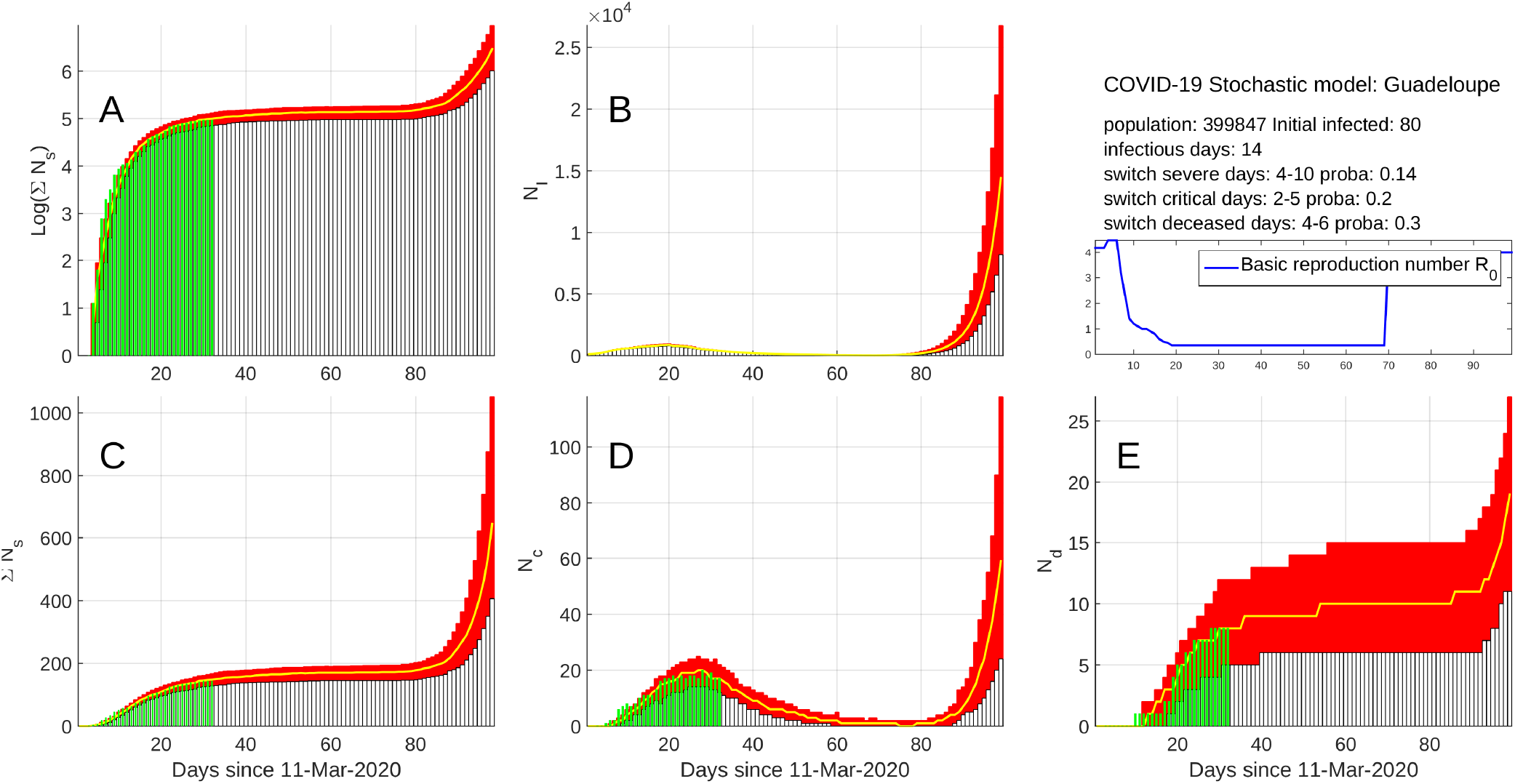
Effects of uncontrolled end of containment. Same model as in Figure 2 but with an uncontrolled end of containment at day 70 with a sudden resetting of *R*_0_ = 4.0. The model indicates that, after approximately one month of low-level infectious spread, the number of cases again dramatically increases after day 90.

The first version of the article was made released on April 16 2020. A third appendix Posterior model assessment with data acquired after April 11, 2020 was added in a second version of the article where the model of Figure 2 is compared to more recent data from April 12 to May 25, 2020 (Fig. 10).

**Fig. 6.**
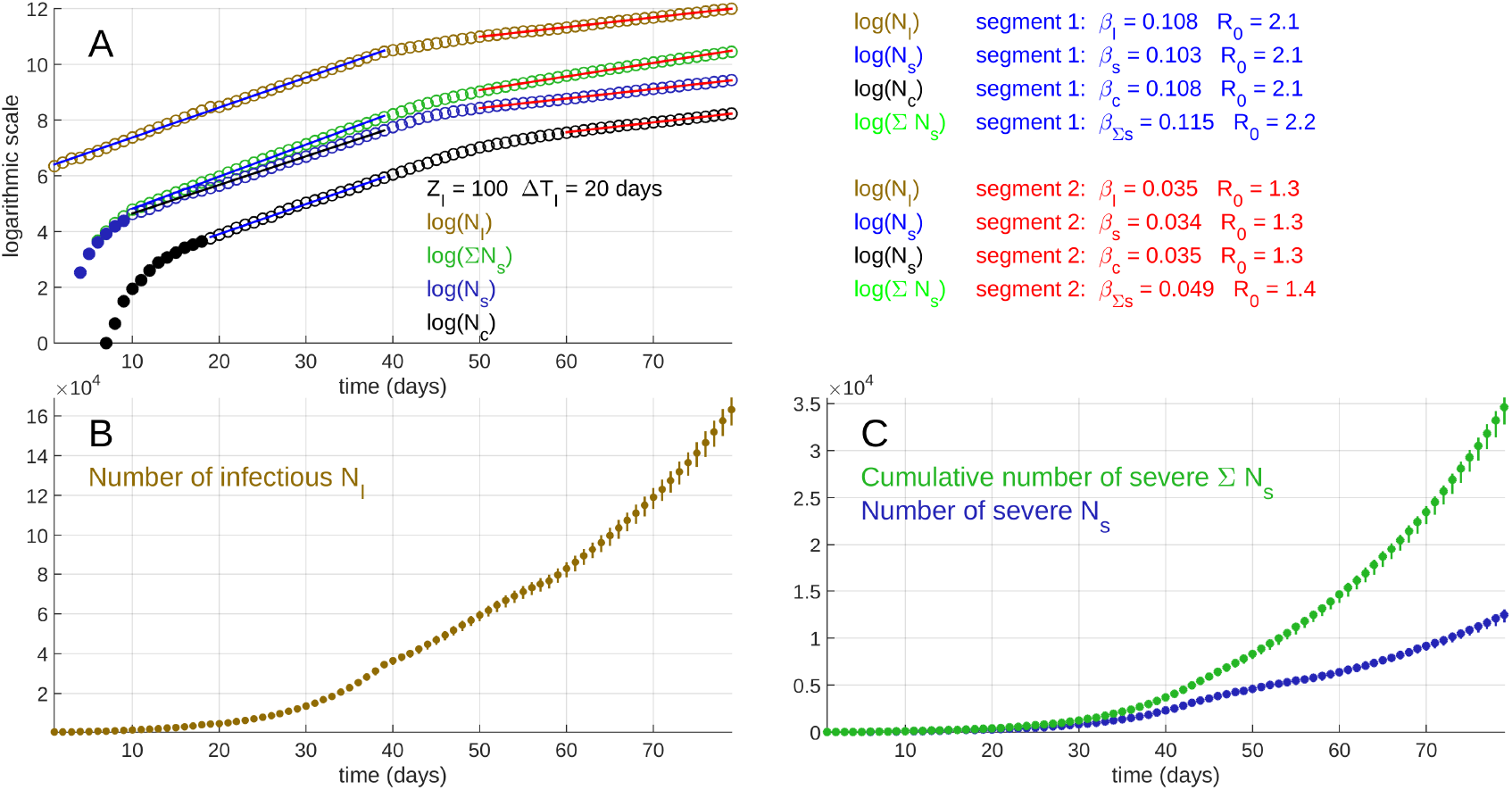
Stochastic simulation for a period of 80 days. B) time variations of *N*_*I*_. C) time variations of Σ*N*_*s*_ and *N*_*s*_ (D). A) The same curves together with *N*_*c*_ displayed in a semi-logarithmic graph (Natural logarithm). The upper right panel of the figure shows the parameters *β* obtained for the 8 linear segments identified in the curves shown in (A). The *R*_0_ values have been derived from *β* using equation 2 with *T*_*I*_ = 14 days. The model started with *Z*_*I*_ = 500 initial infectious “I” and *R*_0_ = 2.0. At day 40, the basic reproduction number changed to *R*_0_ = 1.2. During all the process, the other parameters remained unchanged: *p*_*s*_ = 0.14, *p*_*c*_ = 0.25, *p*_*d*_ = 0.20, Δ*T*_*I*_ = 20 days, Δ*T*_*s*_ = 14 days, Δ*T*_*c*_ = 21 days, and the switch periods *δT*_*s*_ = [4 − 10] days, *δT*_*c*_ = [3 − 9] days, *δT*_*d*_ = [3 − 8] days.

**Fig. 7.**
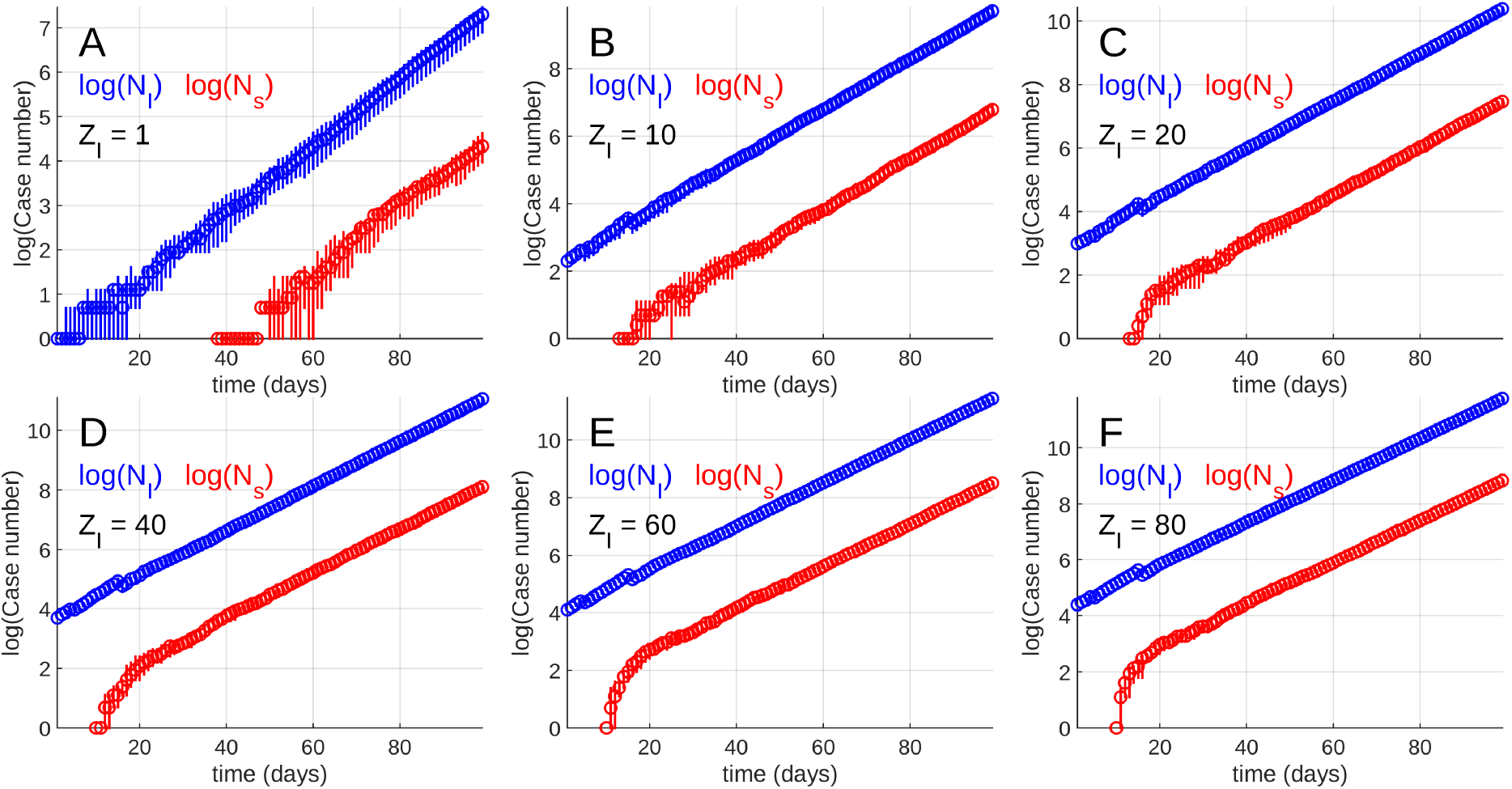
Stochastic simulations obtained for 6 different values of the initial number of infectious. The value of *Z*_*I*_ is indicated on each subplot.

**Fig. 8.**
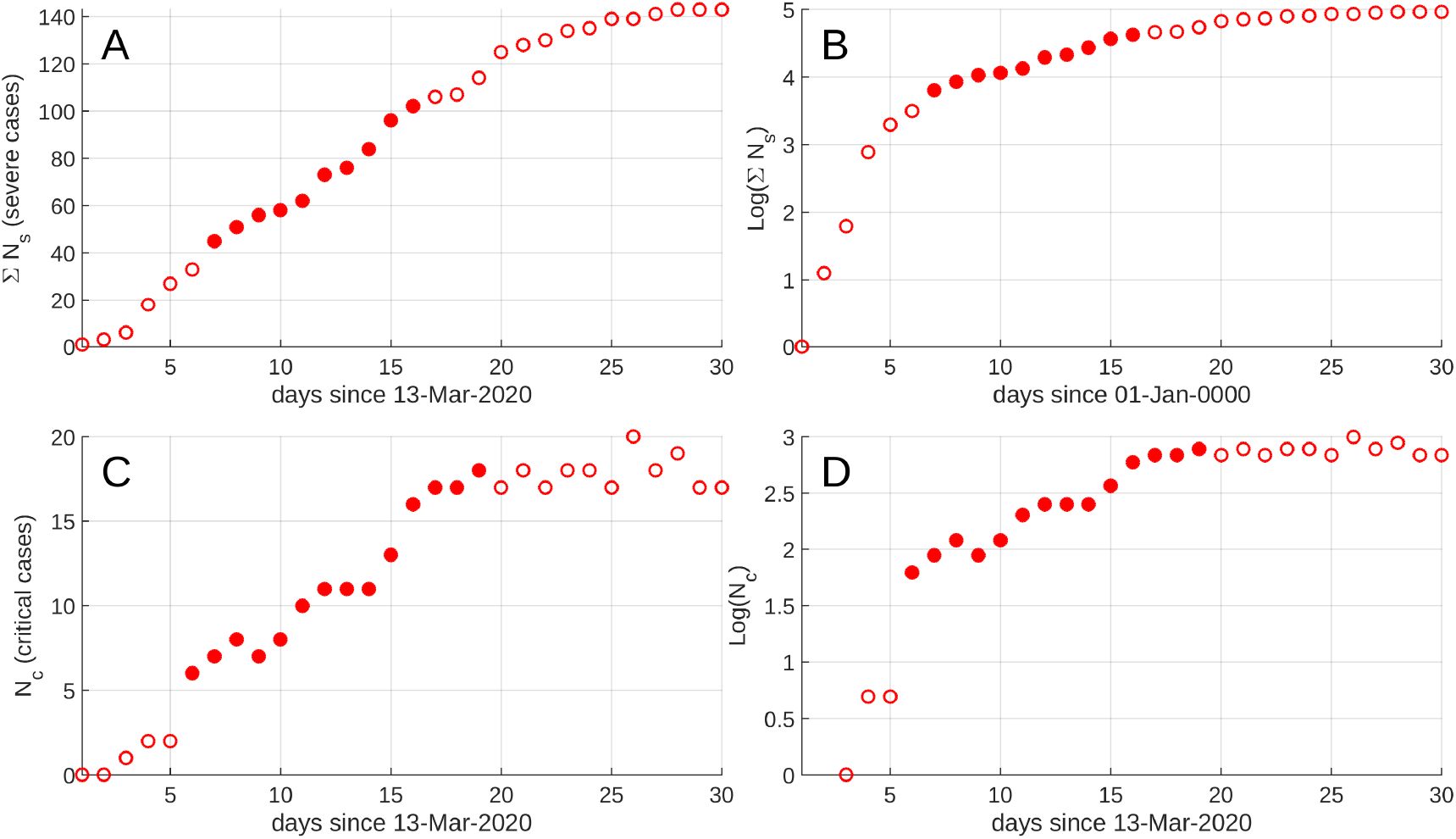
Data used in the present study. Number Σ*N*_*s*_ of severe cases (A) and *N*_*c*_ of critical cases (C) as a function of time. In plots B and D, the data are presented in a semi-logarithmic graph. The filled symbols represent the data points used in the bootstrap computations and are assumed to belong to the linear part of the semi-logarithmic curves when an exponential regime is established.

**Fig. 9.**
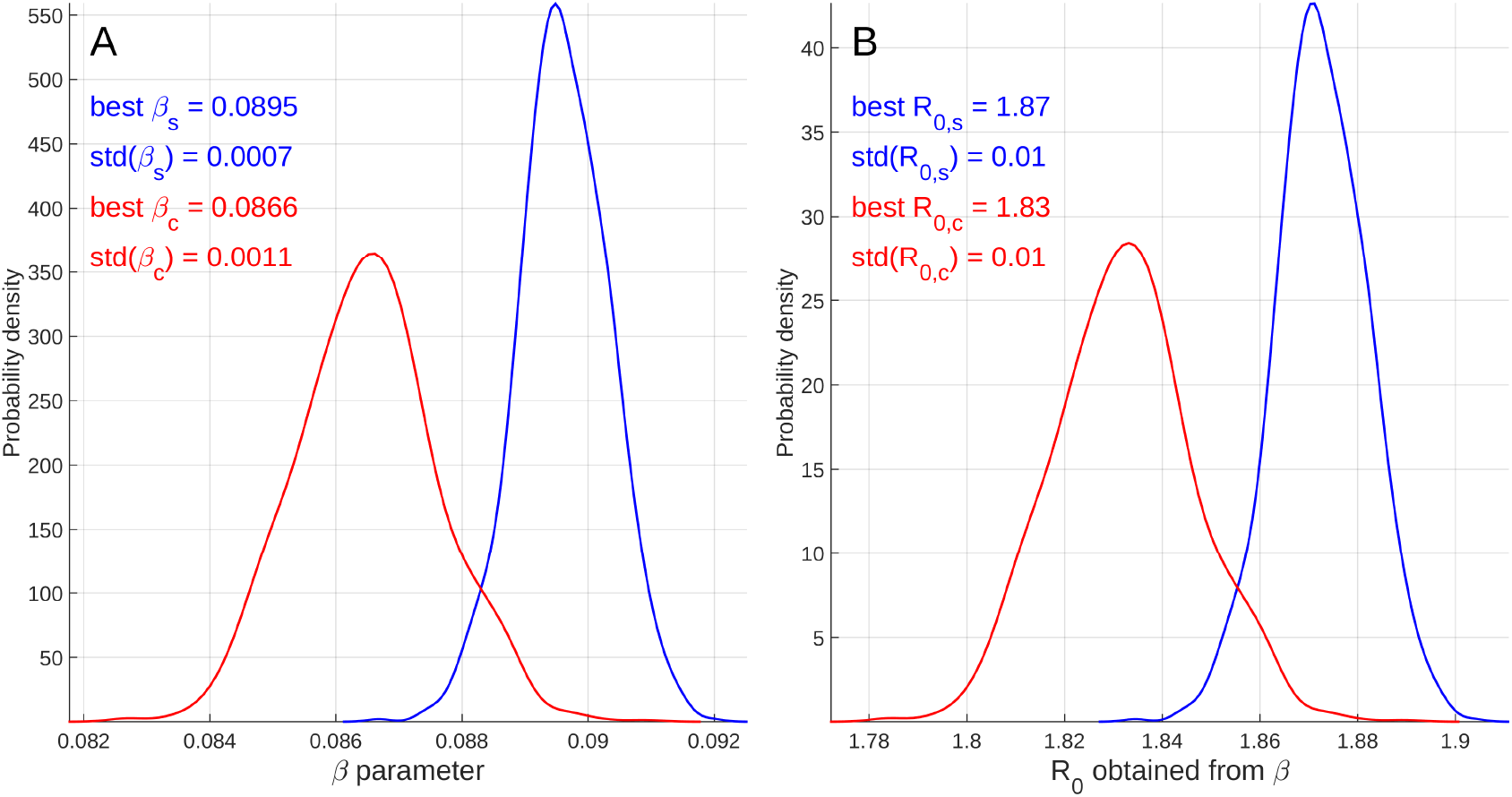
A) Probability density kernels for the parameters *β*_*s*_ and *β*_*c*_ obtained by respectively bootstrapping the data Σ*N*_*s*_ and *N*_*c*_. The data used are represented as filled symbols in Figure 8. B) Kernels for *R*_0_ obtained by applying equation 2 to the bootstrapped parameters *β*_*s*_ and *β*_*c*_.

**Fig. 10.**
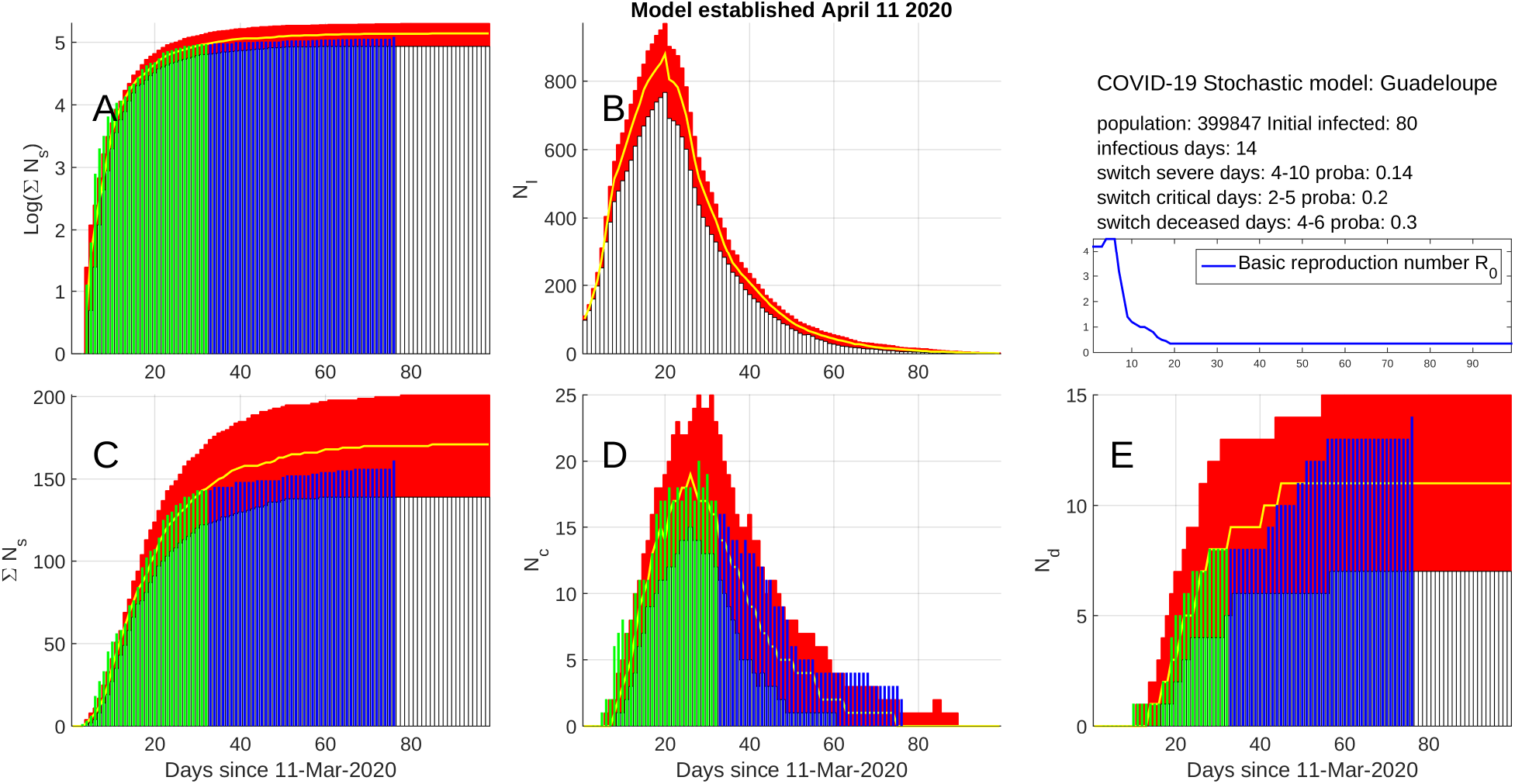
Same Model 1 as in Figure 2 but with the data acquired from April 12 to May 25 2020 (vertical blue bars), i.e. not used to constrain the model.

Figure 10 shows that our predictive model remained particularly accurate during the whole period of COVID-19 decrease, i.e. for a duration of more than 6 weeks. This very good performance of the model may result from the conjunction of several positive factors discussed in appendix Posterior model assessment with data acquired after April 11, 2020. We believe that the most important factors are: i) the design of the model and the data used to constrain the parameters were validated by physicians (MA, PC and MC) belonging to the local University Hospital where all COVID-19 cases of Guadeloupe were treated. This point is crucial to obtain reliable data as emphasised by Sperrin et al. (20). ii) the early confinement made the Guadeloupe archipelago a closed system with no input/output flux of people susceptible to either import or export COVID-19. This made the situation stable and easier to model.

## Data Availability

Data are made publicly available by Sante Publique France.

## Supplementary Note 1: Stochastic Monte Carlo model

### A. Description of the model structure

The stochastic model used in the present study falls in the class of exact stochastic Monte Carlo models (2). In such models, the entire population exposed to an infection is represented by a network where each node represents a person. This type of models offers very large possibilities to design epidemic processes that agree with the medical knowledge of the virus dissemination and its possible pathological issues.

A set of stochastic rules determines the evolution of each node at each time iteration. The infection simulation process begins by choosing a given number of nodes initialised as “infected” to put the pathogen in the network. Other initial conditions can be imposed to fix the state of the population. For instance, we may fix the number of vaccinated persons. In the case of COVID-19 for which no vaccine exists at the present time, the main initial condition is to fix the number *Z*_*I*_ of initially infected individuals. The nest phase of the modelling is a loop over the time steps covering the period to simulate. For each time-step, the state of each node is eventually modified depending on its present stage and according to the set of stochastic rules that define the way by which the infectious disease propagates (a flowchart of the stochastic process is shown in Figure 1).

In our model, we define 6 different possible states for the nodes (Fig. 1):

1. **state “S” for susceptible** corresponds to non-infected people and likely to become infected. In this sense, state “S” is also equivalent to the commonly defined “exposed” state. We note *N*_*S*_ instantaneous number of “S” persons.
2. **state “I” for infectious** is for people infected by COVID-19 and presenting either no or only minor symptoms. These persons are likely to remain undetected by the medical services and expected to pursue their daily activities and maintain contacts with other people. By this way, they are the primary cause of infection diffusion among the set of “S” nodes. The basic reproductive number, *R*_0_, defined as the likely number of “S” infected by a single “I” case during his infectious period *T*_*I*_. We note *N*_*I*_ the instantaneous number of “I” patients.
3. **state “s” for severe** is for people with enough severe symptoms to either see an urban doctor or be admitted in a hospital. These patients are considered to be isolated either at home or at the hospital and are no more able to infect other people. We note *N*_*s*_ the instantaneous number of “s” patients and Σ*N*_*s*_ the corresponding cumulative number.
4. **state “c” for critical** is for patients in a critical state and necessitating intensive care in hospitals. As for “c”, these patients are considered isolated from the “S” population and unable to infect others. We note *N*_*c*_ the instantaneous number of “c” patients.
5. **state “R” for recovered** is for patients “I”, “s” or “c” that recovered after a period of time that depends on the considered state. The recovery periods will be respectively written Δ*T*_*I*_, Δ*T*_*s*_ and Δ*T*_*c*_ for states “I”, “s” and “c”. In the specific case of COVID-19, the main medical opinion is that “R” persons are protected against a new infection by the virus. We note Σ*N*_*R*_ the cumulative number of “R” patients.
6. **state “d” for deceased** patients. We note Σ*N*_*d*_ the cumulative number of “d” patients.

A stochastic set of rules determines the probability to switch from one state to another. In our model, these rules are (Fig. 1):

1. **rule** *S* → *I* **determines the condition to switch from non-infected to infected**. The main parameters of this rule are the infectious period *T*_*I*_ and the basic reproductive number *R*_0_. In our model, this rule is applied to each new “I” node, i.e. nodes that were “S” one day before. For such new “I”, an average number of *R*_0_ are randomly taken among the “S” persons and are randomly set in state “I” in the next *T*_*I*_ days.
2. **rule** *I* → *s* **determines the conditions to switch from infectious to state severe**. This is controlled by a probability level *p*_*s*_.
3. **rule** *s* → *c* **determines the conditions for a patient with severe symptoms to become critical** and will be admitted in a critical care unit. This is controlled by a probability level *p*_*c*_.
4. **rule** *c* → *d* **determines the conditions for a critical patient to die**. This is controlled by a probability level *p*_*d*_.
5. **rule** ∗ → *R* **represents the switch to state “recovered”**. This transition applies to “I”, “s” and “c” states with probability 1 as long as the patients respectively remained in their state for a duration of Δ*T*_*I*_, Δ*T*_*s*_ and Δ*T*_*c*_.

The explicit definition of the rules and the fact that they apply to each node of the network provides a great flexibility to account for more or less sophisticated conditions. For instance, the switching probabilities may easily account for the age of each person. Also, and indeed the model does it, we may consider that a switch from one state to another takes place in a given time interval whose duration is constrained by clinical data. The model is also able to use a time-varying basic reproductive number *R*_0_(*t*_*k*_) in order to account for the effects of containment and social isolation. The nodes may be assigned to different subsets in order to define regions with given populations. Rules may be defined to account for interactions between regions. In the present study, this possibility has not been implemented due to the lack of data to constrain the process.

### B. Model parameters

The model is totally determined by the following parameters:

1. the total number *N* of nodes, i.e. of persons forming the population. For Guadeloupe, we set *N* = 399847 from the age distribution (21).
2. the number *Z*_*I*_ of initial “I”.
3. the basic reproductive number, *R*_0_. This parameter may be time-varying in order to account for different social behaviours. It is generally assumed that *R*_0_ is large for COVID-19, and the range of possible values is large (22). In the present study, we experimentally determine the value of *R*_0_ that best reproduces the observed data. This point will be discussed in details in section Bootstrapping method of data analysis.
4. the infectious period *T*_*I*_ is typically assumed to be of the order of 20 days with possible values as large as 37 days in some exceptional circumstances. In the present study, we determine a value for *T*_*I*_ that best matches with both the data and the prior assumptions taken other studies. This point is considered in section Bootstrapping method of data analysis.
5. the recovery periods Δ*T*_*I*_, Δ*T*_*s*_ and Δ*T*_*c*_ are constrained by clinical data.
6. the switching probabilities *p*_*s*_, *p*_*c*_, and *p*_*d*_ are constrained by clinical data. These probabilities are completed by switch periods, *δT*_*s*_, *δT*_*c*_ and *δT*_*d*_ during which a given state “I”, “s” and “c” may respectively switch to “s”, “c” and “d”.

### C. Simulation examples

In this section we present several simulations to illustrate the effects of the key parameters of the model. This will help the reader to understand where information able to put constrains on the parameters can be obtained from the data processed in section Bootstrapping method of data analysis. In order to quantify the random fluctuations due to the stochastic nature of the model, each simulation is performed 20 times to compute the median and the confidence intervals of the results.

The first simulation corresponds to a duration of 80 days with a basic reproduction number *R*_0_ = 2.0 from day 1 to day 39, and *R*_0_ = 1.2 afterwards. The model started with *Z*_*I*_ = 500 initial infected “I” and the duration of the infectious period is fixed to *T*_*I*_ = 14 days. All other parameters are kept fixed during the process: *p*_*s*_ = 0.14, *p*_*c*_ = 0.25, *p*_*d*_ = 0.20, Δ*T*_*I*_ = 20 days, Δ*T*_*s*_ = 14 days, Δ*T*_*c*_ = 21 days, and the switch periods *δT*_*s*_ = [4−10] days, *δT*_*c*_ = [3−9] days, *δT*_*d*_ = [3−8] days.

Figure 6 shows the results of the simulation for the time variations of *N*_*I*_ (Fig. 6B), and of Σ*N*_*s*_ and *N*_*s*_ (Fig. 6C). The three curves together with *N*_*c*_ are represented in a common semi-logarithmic graph in Figure 6A. The Natural logarithm is used throughout the present paper.

As a starting point for the discussion, it can first be observed that the time variations of *N*_*I*_ and *N*_*s*_ significantly C Simulation examples depart from a pure exponential pattern, particularly because of the presence of smooth bumps in the curves around day 55. These bumps can be better understood in the semilogarithmic plot of Figure 6A where the *N*_*I*_ and *N*_*s*_ curves appear partly linear in two segments. The same linear segments are also visible in the *N*_*c*_ curve. The presence of linear segment in the curves indicates an exponential time variation. In the *N*_*I*_ curve (orange symbols), a first linear segment goes from day 1 until day 39 with slope *β* = 0.108. A second linear segment with slope *β* = 0.035 starts at day 50. A curved segment locates in between the two linear segments, from day 40 to day 50.

The slopes *β* of the linear segments are related to the basic reproductive number through,

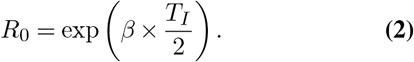

In the present example, taking *T*_*I*_ = 14 days, we have *R*_0_ ≃ 2.1 for the first segment of *N*_*I*_, *N*_*s*_ and *N*_*c*_. For the second segment, *R*_0_ ≃ 1.3 for curves *N*_*I*_, *N*_*s*_ and *N*_*c*_. These values agree very well with the input values used in the model. The linear segment of *N*_*I*_ is slightly biased by the presence of a small jump at day 20 that corresponds to the recovery time Δ*T*_*I*_ = 20 days of the initially infected patients (i.e. 500 persons in this simulation). This cohort of initial infectious massively contaminates *Z*_*I*_*×R*_0_ = 1000 persons, some of these initials switched to state “s” but most of them (i.e. *Z*_*I*_ *×* (1−*p*_*s*_) = 430) recovered and suddenly switched to state “R” at day 20. This produces a sharp decrease of the instantaneous number of infectious patients in the *N*_*I*_ curve. This jump is transmitted in the other curves but highly blurred by the switching process (through stochastic causal convolutions).

The parameter *β* is a primary quantum of information that can be obtained from the data, and equation 2 shows that the parameters *R*_0_ and *T*_*I*_ are linked:

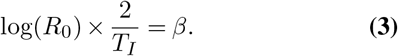

This equation shows that, if *β* is the only information available, the pairs of parameters [*R*_0_, *T*_*I*_] cannot be determined uniquely unless additional information is available through the knowledge of either *R*_0_ or *T*_*I*_.

Information about *T*_*I*_ can be obtained by recognising that this period of time corresponds to the duration of the smooth curved segment that separates the two linear segments discussed above. In curve *N*_*I*_, the curved segment starts at day 40 when the change of *R*_0_ occurs. However, because new infectious patients do not immediately contaminate others but instead do that during the period of time *T*_*I*_, an abrupt change of *R*_0_ appears smoothed. Consequently, this is only after day 50, that a linear segment corresponding to the new value of *R*_0_ appears in the *N*_*I*_ curve. This phenomena is of a considerable practical importance because it represents a latency (or an inertia) of the control measures taken by the authorities to reduce and extinct the epidemic process. Such a latency has to be clearly explained to the population in order to encourage people to maintain they efforts to remain in containment. The two linear segments of the *N*_*s*_ curve (Fig. 6A) are delayed by 9 days with respect to the segments of the *N*_*I*_ curve. This duration of 9 days corresponds to the onset period of the *N*_*s*_ curve during days 1 to 9 of the process (filled blue dots in Fig. 6A). The delay of 9 days is caused by existence of the time period *δT*_*s*_ = [*ts*_1_, *ts*_2_] during which a “I” patient is able to become “s” (in this simulation, *δT*_*s*_ runs from day 4 to day 10). Consequently, the first “s” patients begin to appear after a delay of *ts*_1_ days (i.e. 4 days in this example) and all “s” patients are created at day *t*2_*s*_10. This explains the duration and the shape of the onset period visible at the beginning of the *N*_*s*_ curve. Consequently, the onset period of the *N*_*s*_ curve may provide information about the switch period *δT*_*s*_. The same onset phenomena is observed in the *N*_*c*_ curve but with a delay equals to the sum of *ts*_1_ + *tc*1 = 7 days. The end of the onset period falls at day *ts*_2_ + *tc*_2_ = 19.

We now turn to the case of the Σ*N*_*s*_ curve (green circles in Fig. 6A) which is particularly important because it generally corresponds to the available data. Contrarily to the instantaneous quantities *N*_*I*_ and *N*_*s*_ which give the number of either “I” or “s” patient at a given time, Σ*N*_*s*_ is a cumulative quantity which gives the total number of patients who passed by stage “s” anytime before present. We emphasise that this quantity is NOT the integral of *N*_*s*_ and, as a consequence, the slopes of the linear segments present in the Σ*N*_*s*_ curve are not simply related to those of the *N*_*s*_ curve. Indeed, a careful examination of the Σ*N*_*s*_ reveals that the segments are not strictly linear. At the beginning of the process, we have Σ*N*_*s*_ = *N*_*s*_ until the end of the time periods where first “s” patients begin to switch either to the state “R” or “c”. At that time, the two curves begin to diverge. The slopes of the linear segments in Σ*N*_*s*_ are always slightly larger than the slopes of *N*_*s*_ and the formula 2 and 3 are no more exact for the Σ*N*_*s*_ case. Indeed, the *R*_0_ values derived for Σ*N*_*s*_ in the example (upper right part of Fig. 6) are significantly biased, and to obtain reliable *R*_0_ estimates, it is necessary to use data at the very end of the process, in the narrow time-window comprised between the end of the onset period and the beginning of the switching from “s” to “c”.

The size of the confidence intervals appears constant in the semi-logarithmic plots (Fig. 6A). This is typical of a multiplicative noise where the amplitude of the statistical fluctuations is proportional to the data amplitude as can be checked in Figure 6B,D. In our case, this multiplicative noise may be explained by the growing Brownian divergence of some random walks in the network. Practically, this conducts to the appearance of some outlier simulations and justifies the use of the median.

We now address another important characteristic of the epidemic process through the random variations occurring at the very beginning of the process. The features we want to discuss are illustrated in Figure 7 where the plots have been obtained by running the model with a different number on initial infected *Z*_*I*_. In the case of rather small values of *Z*_*I*_ (i.e. 1, 10 or 20 in Fig. 7A,B,C), random fluctuations perturb the beginning of the curves, with a longer persistence for the *N*_*s*_ curve. For larger values of *Z*_*I*_ (i.e. 40, 60 or 80 in Fig. 7D,E,F), the random fluctuations almost disappear while the starting sequence becomes steeper. Consequently, a careful observation of the starting sequence may provide some information about the number *Z*_*I*_ of initial infectious persons. Let us remark that these features can only be obtained with a stochastic model as the one developed in the present study.

## Supplementary Note 2: Bootstrapping method of data analysis

### A. Presentation of the data

This section gives details on the method used for the data analysis and presents the results used to constrain the stochastic model presented in section Stochastic Monte Carlo model.

Figure 8 shows the data used in the present study, namely the numbers *N*_*s*_ and *N*_*c*_ of severe and critical case observed in Guadeloupe from March 13^*th*^ until April 2 of year 2020. These data are presented in both linear and semi-logarithmic plots in order to better emphasise a possible exponential-like pattern. Because of the small number of data available at the time of writing the present paper, the exponential increase of either *N*_*s*_ or *N*_*c*_ is not as conspicuous as for the synthetic case presented in Figure 6. However, for the data set *N*_*s*_, the semi-logarithmic plot (Fig. 8B) is reasonably linear in the [day 7 − day 16] period. The first 6 points are expected to correspond to the *δT*_*s*_ onset period of 6 to 7 days. For the *N*_*c*_ data, a linear segment may be identified in the [day 6 − day 19] period.

By comparing the onset period in the data with the simulation results of Figure 7, we may claim that the onset sequence of the data curve *N*_*s*_ corresponds to a rather large number of at least 80 initial infectious persons. These persons could for instance be passengers of an aircraft or members of a group infected by a single infectious during a meeting.

### B. Data bootstrapping and parameter determination

In order to determine the *β* parameter and its uncertainty limits from the small-size data sets of Figure 8, we use a bootstrapping approach (23). Let us recall that this method relies on a statistical resampling of the data sets in order to reconstitute the statistical variability of the estimated parameter *β*. In the present study, we performed 1000 bootstrap resamplings for each data set *N*_*s*_ and *N*_*c*_, and the so-obtained 1000 estimates of *β* may be used to compute the probability density kernels shown in Figure 9A. The two probability distributions are poorly statistically coherent with a small overlap of the two curves. Equation 2 may be used to compute *R*_0_ (using *T*_*I*_ = 14 days) from the *β* probability curves. The estimate for *R*_0_ ≃ 1.85 *±* 0.03 is coherent with the values published by Li et al. (22) who found *R*_0_ = 2.2 with a 95% confidence interval [1.4 − 3.9].

## Supplementary Note 3: Posterior model assessment with data acquired after April 11, 2020

The present Section was added in version 2 of the paper to compare the data published after April 11 2020 to the predictions of the model published in the first version of the article. The results are shown in Figure 10 where the data used to constrain the model in version 1 of the article are shown as red vertical bars, and the new data (from April 12 to May 25, 2020), neither used to improve or update the model, are shown as blue vertical bars. It is worth mentioning that the model shown in Figure 10 has been run with the same parameters as those of the “best model” of Figure 2 already published in version 1 of the article. Slight discrepancies between the model results of Figures 2 and 10 are attributable to random fluctuations due to the stochastic nature of the algorithm.

As can be observed in Figure 10, there is a remarkably good agreement between the model predictions and the new data which span a time range of more than 6 weeks. This posterior assessment of our model indicates that modelling the covid19 spread in Guadeloupe with a stochastic approach, although constrained with a relatively small number of parameters, was reliable. This result contradicts repeated claims reporting that the predictions of COVID-19 models are unreliable and should not be used as decision tools (e.g. (24)). Such a negative statement is clearly at odd for the model proposed in the present study and it is interesting to examine why our model has good performances when compared with the low level of performance usually assigned to COVID-19 predictive models.

1. It is worth mentioning that 3 (MA, PC, MC) among the 4 co-authors of the present study are practitioner physicians of the local University Hospital which was the focal point for COVID-19 cares in Guadeloupe. This point is clearly a mandatory condition to obtain reliable data and put pertinent constrains on the model as emphasised by Sperrin et al. (20).
2. The fourth co-author (DG), although not permanently leaving in Guadeloupe, frequently leaves in Guadeloupe since 20 years and has a good knowledge of the social structure of the population.
3. Items 1 and 2 allowed to optimally choose the numerical approach for the Guadeloupe model elaboration:
  a. At the time of the model construction, the input of new information concerning the COVID-19 disease was frequent, abundant and puzzling. The physician co-authors evaluated this amount of information and selected the information to integrate in the model.
  b. The necessity to progressively integrate new and possibly non-linear constrains in the model favoured the choice for an explicit stochastic approach.
4. Specific circumstances concerning the COVID-19 spread in Guadeloupe certainly concurred to the success of our model:
  a. Despite the fact that Guadeloupe is an archipelago, most of the population stays on the Grande Terre and the Basse Terre islands separated by the Rivière Salée with a width of some tens of metres. The industrial, commercial and economic structure of Guadeloupe favours numerous social contacts among the population which can be considered homogeneous with respect to COVID-19 spread.
  b. The spread of COVID-19 in Guadeloupe started several weeks after the spread in Europe and, particularly, in France. For this reason, the containment began a few days after the onset of the COVID-19 spread in Guadeloupe. This probably created a very stable situation during the whole duration of the containment, making the spread more predictable.
  c. Another important particularity of Guadeloupe is that medical and administrative authorities made a very tight control of passengers arriving in Guadeloupe either by plane of by boat. This made the archipelago a closed territory easier to model than open areas.
  d. A coherent and documented communication procedure driven by both the medical and administrative authorities enabled to broadcast convincing and understandable messages to the population. This was reinforced by the fact that these messages were given by respected local physicians with an accurate knowledge of the local sanitary situation.

